# Diabetes and glycemic control as predictors of cardiovascular disease risk biomarkers among people with the metabolic syndrome: a longitudinal analysis

**DOI:** 10.64898/2026.07.29.26359284

**Authors:** Ines Gonzalez-Casanova, Yu Zhou, Estefania Toledo, Dora Romaguera, Angel M Alonso-Gomez, Miguel Fiol, Leire Goicolea-Güemez, Miguel Ángel Martínez-González, Cristina Razquin, Jordi Salas-Salvadó, Montserrat Fitó, Alvaro Alonso

**Affiliations:** Department of Applied Health Science, Indiana University School of Public Health Bloomington, Bloomington, IN, USA; PhD Program in Nutrition, Department of Applied Health Science, Indiana University School of Public Health Bloomington, Bloomington IN, USA; Centro de Investigación Biomédica en Red Fisiopatología de La Obesidad y Nutrición (CIBEROBN), Instituto de Salud Carlos III, Madrid 28029, Spain; Department of Preventive Medicine, School of Medicine, Institute of Nutrition and Health, University of Navarra, IdiSNA, Pamplona, Spain; Health Research Institute of the Balearic Islands (IdISBa), Palma, Spain; Bioaraba Health Research Institute, Osakidetza Basque Health Service, Araba. University Hospital, University of the Basque Country UPV/EHU, Vitoria-Gasteiz, Spain; Universitat Rovira i Virgili, Departament de Bioquímica i Biotecnologia, Alimentació, Nutrició, Desenvolupament i Salut Mental ANUT-DSM, Reus 43204, Spain; Institut de Recerca Biomèdica Catalunya Sud (IISPV), Grup Alimentació, Nutrició, Desenvolupament i Salut Mental, Reus 43204, Spain; Hospital del Mar Research Institute (HMIM), Barcelona, Spain; Department of Epidemiology, Rollins School of Public Health, Emory University, Atlanta, GA, USA

## Abstract

**Background:** Cardiovascular disease (CVD) is a leading cause of morbidity and mortality among individuals with diabetes, where risk remains elevated even with good glycemic control. Cardiac biomarkers such as N-terminal pro-B-type natriuretic peptide (NT-proBNP), high-sensitivity troponin T (hsTnT), high sensitivity C Reactive Protein (hsCRP), Procollagen Type I Carboxy-Terminal Propeptide (PICP), and 3-Nytrotirosine (3-NT) may capture subclinical cardiac stress and injury beyond traditional risk measures.

**Methods:** We analyzed 562 participants from the PREDIMED-Plus trial with data on diabetes status, glycosylated hemoglobin (HbA1c), and cardiovascular biomarkers. At baseline, participants were categorized as having normoglycemia (HbA1c <5.7%; n=50), prediabetes (HbA1c ≥5.7 to <6.5%; n=353), diabetes with well-controlled glycemia (HbA1c <7%; n=127), or diabetes with poorly controlled glycemia (HbA1c ≥7%; n=32). Cardiac biomarkers assessed at baseline, 3 years, and 5 years included NT-proBNP, hsTnT, hsCRP, PICP, and 3-NT. Mixed models, adjusted for demographic, clinical, and lifestyle covariates, were used to examine cross-sectional and longitudinal associations.

**Results:** Cross-sectionally, higher HbA1c was inversely associated with NT-proBNP (β = –0.16, 95% CI: –0.29, –0.04) and directly associated with hsTnT (β = 0.08, 95% CI: 0.02, 0.14), particularly among those with poorly controlled diabetes. No consistent associations were observed for hsCRP, PICP, or 3-NT. Longitudinally, baseline differences in diabetes status were not significantly related to biomarker changes. However, compared to normoglycemic participants, those with well-controlled diabetes showed higher increases in hsTnT (β = 0.10, 95% CI: 0.02, 0.19) and PICP (β = 0.14, 95% CI: 0.01, 0.28).

**Conclusion:** Even with adequate glycemic control, individuals with diabetes may experience progression of subclinical cardiac damage and fibrosis. Incorporating cardiac biomarkers into risk assessment may improve early identification of CVD risk in high-risk populations.

## Introduction

Cardiovascular disease (CVD) remains the leading cause of death globally, accounting for more than 17 million deaths annually.^1^ Individuals with diabetes have a 2 to 4 times greater risk of CVD compared to their counterparts without diabetes.^2, 3^ Potential pathways explaining this higher risk include chronic hyperglycemia, systemic inflammation, and accompanying comorbidities such as hypertension and dyslipidemia.^2^ Glycemic control contributes to CVD prevention in individuals with diabetes; however, patients with well-controlled diabetes continue to experience elevated cardiovascular risk when compared to individuals without diabetes, suggesting that the presence of subclinical disease mechanisms beyond glucose levels alone requires further study.^4^ Elucidating the biomarkers involved in the onset and progression of CVD and the role of glycemic control in these mechanisms can contribute to improve CVD prevention in individuals with and without diabetes.

Cardiac biomarkers such as N-terminal pro-B-type natriuretic peptide (NT-proBNP), high-sensitivity cardiac troponin T (hsTnT), high-sensitivity C-reactive protein (hsCRP), and 3-nitrotyrosine (3-NT) have emerged as potentially sensitive indicators of subclinical cardiovascular stress and injury. For instance, NT-proBNP has been associated with higher risk of death and cardiovascular events in people with pre-diabetes and diabetes independently of glycemic status.^5, 6^ Similarly, a recent study among adults with type 2 diabetes found that a single measurement of hsTnT was strongly associated with CVD risk.^7^ Individuals with diabetes also tend to have higher concentrations of CRP, a marker of inflammation, which is also associated with CVD complications.^8^ Meanwhile, 3-NT is a marker of oxidation and endothelial disfunction and is associated with atherosclerosis in both people with and without diabetes.^9^ Hence, while there is some evidence of the role of these biomarkers as indicators of CVD risk in people with diabetes, the role of glycemic control on the concentration of these biomarkers, especially over time, has not been elucidated yet.

Given the increasing prevalence of diabetes and its silent contribution to CVD burden, there is a growing need to understand how glycemic control interacts with cardiovascular biomarkers to influence disease trajectory. By comparing individuals with and without diabetes, this study aims to evaluate both cross-sectional and longitudinal associations between glycemic status and key cardiac biomarkers to better characterize early cardiovascular risk profiles in this high-risk population.

## Methods

### Study participants

This analysis was conducted in a subsample of PREDIMED-Plus trial participants with information on self-reported or diagnosed diabetes status, glycosylated hemoglobin (HbA1c) and biomarkers associated with cardiovascular risk. The PREDIMED-Plus study is a multicenter, randomized controlled trial designed to assess the effect of an intensive weight-loss intervention alongside an energy-reduced Mediterranean diet and behavioral support on the primary prevention of cardiovascular disease in 6,874 participants with overweight/obesity and metabolic syndrome. The inclusion and exclusion criteria, as well as the trial design, implementation, and impact on diabetes incidence and glycemic outcomes for PREDIMED-Plus (ISRCTN89898870) have been described elsewhere.^10–12^ Briefly, participants were recruited from 23 participating centers in Spain and randomly assigned to an intensive lifestyle intervention based on an energy-reduced Mediterranean diet, increased physical activity, and cognitive-behavioral weight management or to a control intervention of low-intensity dietary advice on the Mediterranean diet (not energy-reduced). Inclusion criteria for the main trial were men aged 55–75 years and women aged 60–75 years, with overweight or obesity (body mass index 27–40 kg/m^2^), who at baseline met at least three components of the metabolic syndrome. ^10^ Exclusion criteria included documented history of cardiovascular disease among others. ^10^ For this analysis, we included a subsample of PREDIMED-Plus participants from three participating centers (University of Navarra, Araba University Hospital, Son Espases University Hospital), which had additional information on relevant atrial fibrillation biomarkers measured at least at one time point (baseline, 3 years, 5 years).

Study protocols were approved by Institutional Review Boards of participating institutions. All participants provided written informed consent (ISRCTN89898870).

### Diabetes Status at Baseline

A variable combining information from participants’ self-report of diabetes status, medication status, and measured HbA1c and fasting glucose at screening and baseline was developed and used to assess diabetes information at baseline. HbA1c levels were categorized into four groups based on diabetes status. Participants without diabetes (and not taking diabetes medication) with HbA1c levels below 5.7% were classified as normoglycemic (non-diabetic), while those with HbA1c levels between 5.7% and 6.5% and not taking diabetes medications were considered as having pre-diabetes. Participants were classified as having diabetes if they met any of the following criteria at baseline: self-reported diabetes diagnosis, HbA1c ≥6.5% (48 mmol/mol), elevated fasting glucose (126 mg/dL (7.0 mmol/L) or higher), or taking antidiabetic medication. Among those identified as having diabetes, glycemic control was further categorized. Well-controlled diabetes was defined as having an HbA1c below 7%, while non-well controlled glycemia was defined as HbA1c ≥7.0%, based on clinical recommendations for glycemic targets.^13^ (Figure 1) No distinction was made between type 1 and type 2 diabetes in this analysis.

**Figure 1:**
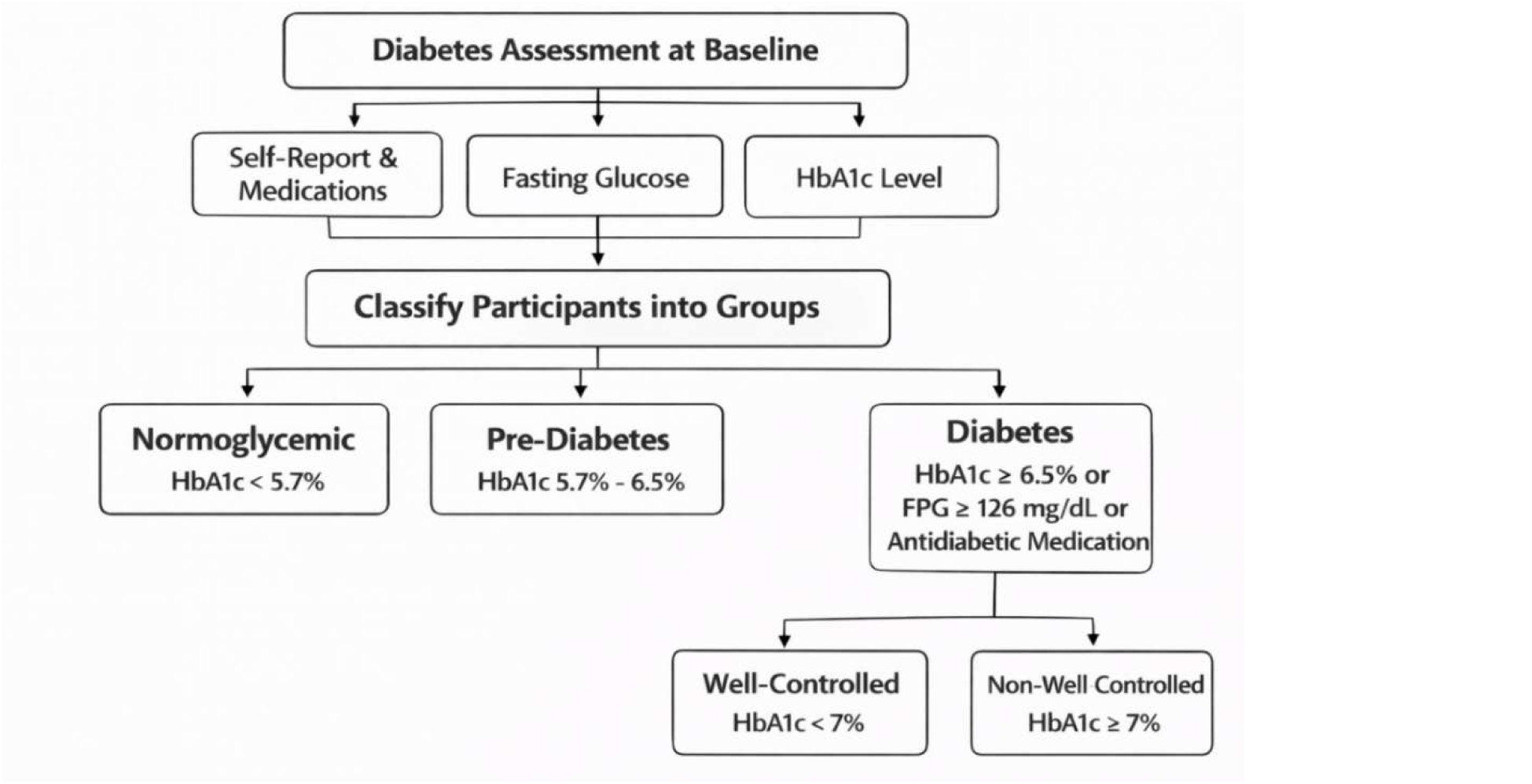
Diabetes classification at baseline

### Biomarker measurements

Glycosylated hemoglobin (HbA1c) was assessed at baseline, 6 months, 12 months, 3 years, 5 years visits from fasting blood samples. Cardiovascular risk biomarkers included in this analysis were NTproBNP and hsTnT, markers of myocardial stress and damage;^5–7^ hsCRP, an indicator of inflammation associated with an increased risk of cardiovascular events, including myocardial infarction and stroke;^8^ 3-NT, a marker of oxidative stress associated with atherosclerosis and endothelial dysfunction; and PICP, a marker of myocardial fibrosis.^9^ These biomarkers were measured from fasting blood samples at baseline, 3-year, and 5-year follow-ups. NTproBNP and hsTnT were measured through electrochemiluminescence immunoassay, while hsCRP was measured through immunoturbidimetry and were run on a Cobas 8000 autoanalyzer (Roche Diagnostics). Enzyme-linked immunosorbent assay technique (ELISA) was utilized to measure 3-NT (Human Nitrotyrosin ELISA kit; Abcam, Cambridge, UK) and PICP (MicroVue PICP EIA; Quidel, San Diego, CA, USA). The intra- and inter-assay variability for biomarkers are as follows: 3.8% and 6.9% for NTproBNP; 2.0% and 3.7% for hsTnT; 3.2% and 3.9% for hsCRP; 5.5% and 7.2% for PICP; and 10% and 15% for 3-NT.

### Covariates

Participants’ sociodemographic and health characteristics were assessed at baseline via a face-to-face questionnaire. Sociodemographic variables included age at baseline, sex, marital status (categorized into married or not married which included single, widowed, and divorced), education (years of schooling), occupation (employed, unemployed, retired, housework). Body mass index was measured at baseline and included in the analysis, as well as self-reported history of diabetes, hypertension, depression, or non-AF arrythmia. The 17-item energy-reduced Mediterranean Diet Adherence Screener was used to assess adherence to the energy-reduced Mediterranean Diet.^14^ The recruitment site (Mallorca, Vitoria, or Navarra) was also included as a covariate in this analysis. Similarly, longitudinal models were adjusted for intervention group based on the impact of the intervention on diabetes and glycemic control outcomes.^12^

### Statistical analysis

We categorized participants into four analytic groups: 1) those without diabetes and normal HbA1c (normoglycemic), 2) those with prediabetes, 3) those with diabetes and well-controlled HbA1c (<7%), and 4) those with diabetes and not well-controlled HbA1c (≥7%). Descriptive analyses were used to assess the different covariates by these groups.

To test for cross-sectional associations between HbA1c (%) – a continuous variable- and diabetes/glycemic control categories with the biomarkers, we used minimally adjusted (age, sex, and education) and fully adjusted (age, sex, education, marital status, smoking, physical activity, height, body mass index, depression, and adherence to the energy-reduced Mediterranean diet) linear regression models.

Similar models were conducted to test associations with biomarker change over time: To assess if diabetes/glycemic control categories at baseline predicted 5-year change in biomarker measurements, we used mixed models predicting the slope along the three time-point measurements (baseline, and 3-year, and 5-year follow ups). Mixed models were also used to assess associations between baseline HbA1c (%) as a continuous variable and 5-year change in biomarkers including 3 time points (baseline, 3 years, 5 years. Similarly, mixed models were used to assess if the slope of HbA1c was associated with 5-year change in biomarkers. Besides the previously mentioned covariates, all longitudinal models were adjusted for intervention arm.

Biomarkers were log-transformed to normalize their distribution for all cross-sectional and longitudinal analyses.

Minimally adjusted and fully adjusted models described above were used to test for all associations. All analyses were conducted using SAS 9.4.

## Results

The final sample for this analysis included 562 participants with available diabetes status, HbA1c and biomarker information. Most of the participants were in the prediabetes group at baseline. Most participants were male, whereas the normoglycemic group had the lowest proportion of female participants (24%).

**Table 1:** Baseline characteristics by baseline diabetes/glycemic control categories.

|  | Normoglycemic | Prediabetes | Well-controlled Diabetes | Not well-controlled Diabetes |
| --- | --- | --- | --- | --- |
| N | 50 | 353 | 127 | 32 |
| Age (years) | 64.7(4.9) | 65.1 (4.9) | 65.4 (4.5) | 65.9 (4.9) |
| Female, % | 24.0 | 41.1 | 39.4 | 50.0 |
| Educational level, % |  |  |  |  |
| College or technical studies | 20.0 | 19.0 | 22.1 | 17.2 |
| High school | 35.6 | 32.9 | 29.4 | 21.9 |
| Less than high school | 44.4 | 48.1 | 48.5 | 60.9 |
| Site, % |  |  |  |  |
| Mallorca | 46.0 | 27.2 | 27.6 | 21.9 |
| Navarra | 30.0 | 21.5 | 22.8 | 21.9 |
| Vitoria | 24.0 | 51.3 | 49.6 | 56.2 |
| Marital status, % |  |  |  |  |
| Single | 6.1 | 5.1 | 7.1 | 6.3 |
| Married | 75.5 | 79.6 | 77.2 | 68.8 |
| Widower | 8.2 | 8.8 | 11.8 | 18.8 |
| Divorced or separated | 10.3 | 6.5 | 3.9 | 6.2 |
| Body mass index (kg/m<sup>2</sup>) | 32.1 (3.0) | 32.2 (3.4) | 32.4 (3.7) | 32.8 (3.4) |
| Hypertension, % | 73.3 | 85.4 | 83.8 | 89.1 |
| Current smoker, % | 4.0 | 10.2 | 9.5 | 9.4 |
| Former smoker, % | 50.0 | 49.9 | 55.1 | 54.7 |
| Mediterranean diet adherence, (0-17 points) | 8.0 (3.2) | 7.4 (2.9) | 7.6 (2.8) | 7.7 (2.4) |
| In the intervention group, % | 58.0 | 48.7 | 48.0 | 50.0 |
| Biomarkers at baseline (median, interquartile range) |  |  |  |  |
| NT-proBNP, pg/mL | 76.1 (102.2) | 95.8 (166.5) | 86.6 (206.5) | 79.8 (133.5) |
| hsTnT, ng/L | 9.3 (3.8) | 9.3 (4.3) | 8.9 (4.2) | 11.3 (8.1) |
| hsCRP, mg/dL | 0.3 (0.4) | 0.4 (0.8) | 0.5 (0.6) | 0.4 (0.5) |
| PICP, mg/mL | 97.8 (41.9) | 97.0 (40.8) | 99.4 (37.9) | 95.9 (52.7) |
| 3-NT, nM | 807.8 (639.1) | 782.1 (755.1) | 746.4 (659.9) | 549.2 (430.0) |

Presented as means (SD) or percentages. NT-proBNP, N-terminal pro-B-type natriuretic protein. hsTnT, high sensitivity troponin-T. hsCRP, high-sensitivity C-reactive protein. 3-NT, 3-nitrotyrosine. PICP, procollagen type 1 carboxy-terminal propeptide.

Table 2 presents cross-sectional associations of diabetes status and HbA1c with concentrations of cardiac biomarkers. At baseline, in multivariate analyses, HbA1c was inversely associated with NT-proBNP (b = −0.16; 95% CI: −0.29, −0.04). Similarly, those participants with uncontrolled diabetes had lower levels of NT-proBNP compared to those with normoglycemia (b = −0.29; 95% CI: −0.56, −0.02) at baseline. In contrast, HbA1c was directly associated with concentrations of hsTnT (b = 0.07; 95% CI: 0.03, 0.11), and participants in the uncontrolled diabetes group had higher concentrations of hsTnT compared to those in the normoglycemic group (b = 0.16, 95% CI: 0.04, 0.29).

**Table 2:** Association of diabetes and HbA1c concentrations with concentrations of log-transformed biomarkers at baseline in a cohort of adults with overweight/obesity and metabolic syndrome.

|  |  | Normoglycemic | Prediabetes | Diabetes well-controlled | Diabetes not well-controlled | Baseline HbA1c (%) |
| --- | --- | --- | --- | --- | --- | --- |
| NT-proBNP | Model 1 | REF | 0.04 (-0.14, 0.23) | -0.10 (-0.36, 0.16) | -0.25 (-0.53, 0.02) | -0.09 (-0.19, 0.01) |
|  | Model 2 | REF | -0.003 (-0.19, 0.18) | -0.11 (-0.38, 0.14) | -0.29 (-0.56, -0.02)* | -0.16 (-0.29, -0.04)* |
| hsTnT | Model 1 | REF | 0.02 (-0.07, 0.10) | -0.02 (-0.13, 0.11) | 0.16 (0.04, 0.29)* | 0.07 (0.03, 0.11)* |
|  | Model 2 | REF | 0.02 (-0.06, 0.11) | -0.02 (-0.14, 0.10) | 0.16 (0.03, 0.28)* | 0.08 (0.02, 0.14)* |
| hsCRP | Model 1 | REF | 0.05 (-0.15, 0.25) | 0.29 (0.01, 0.57)* | 0.10 (-0.20, 0.40) | 0.09 (-0.02, 0.19) |
|  | Model 2 | REF | 0.07 (-0.12, 0.27) | 0.27 (-0.003, 0.55) | 0.07 (-0.22, 0.36) | 0.04 (-0.09, 0.17) |
| PICP | Model 1 | REF | -0.02 (-0.11, 0.07) | -0.02 (-0.15, 0.11) | -0.03 (-0.17, 0.10) | -0.02 (-0.07, 0.03) |
|  | Model 2 | REF | -0.03 (-0.12, 0.06) | -0.02 (-0.15, 0.11) | -0.04 (-0.18, 0.10) | -0.03 (-0.10, 0.03) |
| 3-NT | Model 1 | REF | -0.06 (-0.24, 0.12) | -0.08 (-0.33, 0.18) | -0.30 (-0.57, -0.03)* | -0.09 (-0.19, 0.01) |
|  | Model 2 | REF | -0.03 (-0.21, 0.15) | -0.02 (-0.28, 0.24) | -0.24 (-0.51, 0.03) | -0.06 (-0.19, 0.06) |

*HbA1c, glycated hemoglobin; NT-proBNP, N-terminal pro-B-type natriuretic peptide; hsTnT, high-sensitivity cardiac troponin T; hsCRP, high-sensitivity C-reactive protein; PICP, procollagen type I C-terminal propeptide; 3-NT, 3-nitrotyrosine; REF, reference group. Model 1 was adjusted for prespecified covariates. Model 2 was additionally adjusted for baseline HbA1c. Values are presented as regression coefficients (95% confidence intervals). $P < 0.05$ . Model 1: adjusted for age, sex, research site, and education. Model 2: adjusted for age, sex, research site, education, marital status, smoking, physical activity, height, body mass index, systolic and diastolic blood pressure, depression, and adherence to the energy-reduced Mediterranean diet. Among people with diabetes, those with HbA1c concentrations $<7\%$ were categorized as well-controlled and those with concentrations $\geq 7\%$ were categorized as not-well controlled. \* $p < 0.05$*

In the 5-year longitudinal analysis, no significant associations were observed between HbA1c at baseline and changes in any biomarkers, including NT-proBNP, hsTnT, hsCRP, PICP, or 3-NT. However, 5-year change in HbA1c was inversely associated with NT-proBNP. Similarly, compared to the normoglycemic group, participants with well-controlled diabetes experienced significant increases in hsTnT (Model 1: b = 0.11, 95% CI: 0.03 to 0.20; Model 2: b = 0.10, 95% CI: 0.02 to 0.19) and PICP (Model 1: b = 0.14, 95% CI: 0.01 to 0.28; Model 2: b = 0.15, 95% CI: 0.01 to 0.28). No significant associations were observed for the not well-controlled diabetes group. (Table 3)

**Table 3:** Association of diabetes and HbA1c concentrations with 5-year change in log-transformed biomarkers from baseline through year 5 in a cohort of adults with the metabolic syndrome.

|  |  | Normoglycemic | Prediabetes | Diabetes well-controlled | Diabetes not well-controlled | Baseline HbA1c (%) | HbA1c (%) slope |
| --- | --- | --- | --- | --- | --- | --- | --- |
| NT-proBNP | Model 1 | REF | -0.05 (-0.21, 0.11) | -0.03 (-0.26, 0.20) | 0.09 (-0.15, 0.34) | 0.00 (-0.07, 0.07) | -0.09 (-0.17, -0.01)* |
|  | Model 2 | REF | -0.03 (-0.20, 0.13) | -0.02 (-0.26, 0.21) | 0.08 (-0.16, 0.32) | 0.06 (-0.05, 0.17) | -0.13 (-0.23, -0.03)* |
| hsTnT | Model 1 | REF | 0.02 (-0.04, 0.07) | 0.11 (0.03, 0.20)* | 0.01 (-0.08, 0.10) | 0.02 (-0.01, 0.05) | 0.02 (-0.01, 0.05) |
|  | Model 2 | REF | 0.01 (-0.05, 0.07) | 0.10 (0.02, 0.19)* | -0.002 (-0.09, 0.09) | 0.001 (-0.03, 0.04) | 0.02 (-0.04, 0.07) |
| hsCRP | Model 1 | REF | 0.04 (-0.14, 0.23) | -0.20 (-0.46, 0.07) | 0.16 (-0.11, 0.44) | 0.02 (-0.08, 0.12) | 0.12 (0.03, 0.21)* |
|  | Model 2 | REF | 0.07 (-0.12, 0.26) | -0.19 (-0.46, 0.08) | 0.19 (-0.09, 0.48) | 0.02 (-0.19, 0.24) | 0.12 (-0.04, 0.29) |
| PICP | Model 1 | REF | 0.04 (-0.05, 0.14) | 0.14 (0.01, 0.28)* | -0.02 (-0.17, 0.12) | 0.01 (-0.04, 0.06) | 0.00 (-0.04, 0.05) |
|  | Model 2 | REF | 0.04 (-0.06, 0.13) | 0.15 (0.01, 0.28)* | -0.03 (-0.17, 0.12) | 0.03 (-0.07, 0.14) | 0.01 (-0.07, 0.09) |
| 3-NT | Model 1 | REF | -0.03 (-0.17, 0.12) | 0.003 (-0.20, 0.21) | -0.05 (-0.26, 0.17) | 0.003 (-0.07, 0.08) | -0.04 (-0.12, 0.04) |
|  | Model 2 | REF | -0.03 (-0.18, 0.12) | 0.01 (-0.20, 0.21) | -0.06 (-0.28, 0.15) | 0.08 (-0.10, 0.24) | -0.03 (-0.17, 0.10) |

*HbA1c, glycated hemoglobin; NT-proBNP, N-terminal pro-B-type natriuretic peptide; hsTnT, high-sensitivity cardiac troponin T; hsCRP, high-sensitivity C-reactive protein; PICP, procollagen type I C-terminal propeptide; 3-NT, 3-nitrotyrosine; REF, reference group. Model 1: adjusted for age, sex, and education. Model 2: adjusted for age, sex, education, marital status, smoking, physical activity, height, body mass index, depression, and adherence to the energy-reduced Mediterranean diet, and intervention group. Diabetes well-controlled is defined as HbA1c concentration below 7% for people with diabetes. Diabetes not well-controlled is defined as HbA1c above or equal to 7% for people with diabetes. \*p<0.05.*

The sensitivity analysis excluding people with normoglycemia resulted in similar associations to those generated with the models including the entire sample (Supplemental Table 1).

## Discussion

In this study, we examined both cross-sectional and longitudinal associations between diabetes/glycemic control categories, HbA1c, and biomarkers linked to cardiovascular disease among participants in the PREDIMED-Plus trial. At baseline, HbA1c was inversely associated with NT-proBNP and directly associated with hsTnT, whereas no consistent associations were found for PICP, hsCRP, or 3-NT. Baseline HbA1c was not significantly associated with 5-year changes in any biomarker. However, after categorizing by glycemic control, participants with well-controlled diabetes showed significantly greater increases in hsTnT and PICP compared to their counterparts with normal blood glucose levels.

In this sample of adults with metabolic syndrome, the strongest association between glycemia, diabetes status, and a biomarker was with hsTnT. This biomarker, an indicator of myocardial injury shown to predict adverse cardiovascular outcomes in both diabetic and non-diabetic populations,^15^ was cross-sectionally associated with HbA1c and presented higher levels among participants with uncontrolled diabetes compared to those with normoglycemia. Additionally, hsTnT had a significantly greater 5-year increase among individuals with well-controlled diabetes compared to their counterparts with normoglycemia at baseline. This greater 5-year change was not observed among individuals with prediabetes or those with uncontrolled diabetes at baseline when compared to those with normoglycemia at baseline. These findings from the longitudinal analysis suggest that even when blood glucose is well managed, subclinical cardiovascular changes may continue to progress. In contrast, it is possible that individuals in our study who had poorly controlled diabetes may have already reached persistently elevated hsTnT levels, leaving less room for further increases over time. Beyond its traditional role in detecting acute coronary syndromes, recent evidence suggests hsTnT may also reflect broader microvascular complications of diabetes, including peripheral nerve injury and early cardiomyocyte stress.^16^ These biomarkers offer the potential to identify cardiovascular risk at an earlier stage than traditional clinical measures, particularly in individuals with well-controlled diabetes who may not otherwise be flagged as high risk.^17, 18^

While hsTnT is well known as a marker of subclinical myocardial injury, growing evidence suggests it may also reflect broader microvascular damage linked to chronic high blood glucose. For example, elevated hsTnT levels have been associated with nerve damage in individuals with impaired glucose tolerance and type 2 diabetes, suggesting that this biomarker captures signs of microangiopathy beyond the heart.^16^ These observations support the idea that even when blood glucose is under control, subtle cardiac injury may still progress particularly in patients with the metabolic syndrome, such as participants in our study. Findings from a secondary analysis of the Dapagliflozin Effect on Cardiovascular Events–TIMI 58 (DECLARE-TIMI 58) trial, showed that hsTnT can help identify individuals with type 2 diabetes who are at higher cardiovascular risk and may benefit more from certain therapies, such as SGLT2 inhibitors.^19^ Findings from our analysis support further research to assess the potential of hsTnT as an early marker for cardiovascular risk in diabetes, even in the absence of overt disease.

Other biomarkers had less consistent associations with HbA1c and glycemic status. For instance, NT-proBNP was inversely associated with HbA1c at baseline and with change in HbA1c over time. This biomarker is released in response to myocardial wall stress and volume overload and has been independently associated with future risk of heart failure, coronary events, and stroke, even in individuals without overt symptoms or diagnosed cardiovascular conditions.^8, 20^ However, metabolic factors such as insulin resistance and excess adiposity may suppress natriuretic peptide production, particularly among people with type 2 diabetes,^21^ which could explain this inverse association. Similarly, it is important to highlight the unique nature of participants in this study who have the metabolic syndrome and high-risk for CVD.

On average, PICP, a marker of cardiac fibrosis, had a greater 5-year increase among participants with well-controlled diabetes at baseline compared to those with normoglycemia at baseline. In this sense, PICP has also been proposed as a marker of failing B-cell function and as a good predictor of diabetes disease progression.^22, 23^ The longitudinal associations found among people with well-control diabetes (but not among people with not-well controlled diabetes) could be signaling this progression of the diabetes pathology over time or could be a result of other cardiometabolic risk factors in this population.

These longitudinal findings showing higher increases in PICP only among participants with well controlled diabetes but not among those with not well controlled diabetes at baseline is unexpected, but similar to what was found for hsTnT in this study. It is possible that participants in the not well-controlled group are already at very high levels of these biomarkers and thus there was not a higher increase over time, whereas those in the well-controlled diabetes group still had room for the disease to progress and for these biomarkers to increase. It is also possible that these biomarkers are increasing as a response to other CVD risk factors present in this population beyond diabetes or glycemic control.

There are important limitations to consider in interpreting the results of this study. First, its observational design limits our ability to draw causal conclusions. Second, the relatively small sample sizes in certain subgroups may reduce the statistical power to detect meaningful changes, especially over time. Multiple testing can increase the likelihood of false positive associations; however, we observed a greater number of positive findings than those expected by chance, and the associations observed have biological plausibility and are consistent across models. Finally, despite adjusting for a comprehensive set of covariates, we cannot rule out the possibility of residual confounding or chance findings. Conversely, strengths of this study include the use of longitudinal biomarker data, along with comprehensive adjustment for a wide range of lifestyle, clinical variables, and a panel of circulating cardiac biomarkers. By focusing on both glycemic control and subclinical cardiovascular risk in a high-risk population, the study provides valuable insights into early disease progression. Additionally, distinguishing between well-controlled and poorly controlled diabetes adds important nuance, helping us better understand how cardiovascular risk may vary depending on glycemic control.

In conclusion, in this secondary data analysis of a cohort of participants with the metabolic syndrome, we found higher increases in the concentration of biomarkers of CVD risk - especially hsTnT and PICP - among participants with well-controlled diabetes at baseline compared to those with normoglycemia. These findings could reflect the increased CVD risk of people living with diabetes even under glycemic control.

## Data Availability

Data collaboration for PREDIMED-Plus study is guided by the Data Sharing and Management guide. We follow a controlled data collaboration model, using anonymised (de-identified) study data only, for collaborating with approved researchers.Requests are considered by the PREDIMED-Plus Steering Committee composed by Salas-Salvadó J (Chair), Martínez-González MA, Fitó M. Ros E, Tinahones F, Corella D and Estruch R. Decisions on data access are based on the scientific legitimacy of the requester and of their institution, and on assurances on information security and governance; and with regard to the study's scientific reputation, the needs of funded study team research, the terms of participant consent, and regulatory requirements.

https://www.predimedplus.com/en/project/

---

**Supplemental Table S1.**
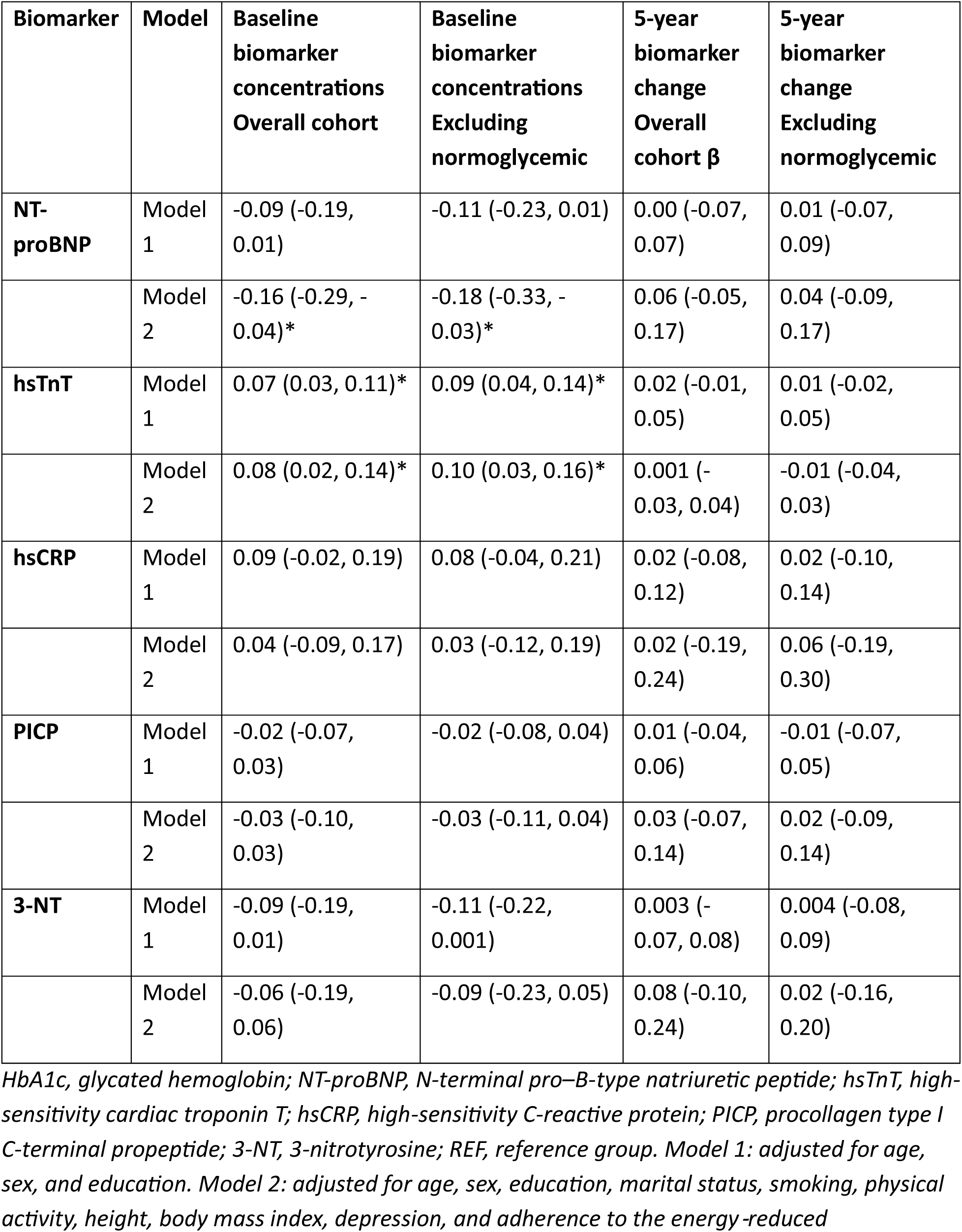
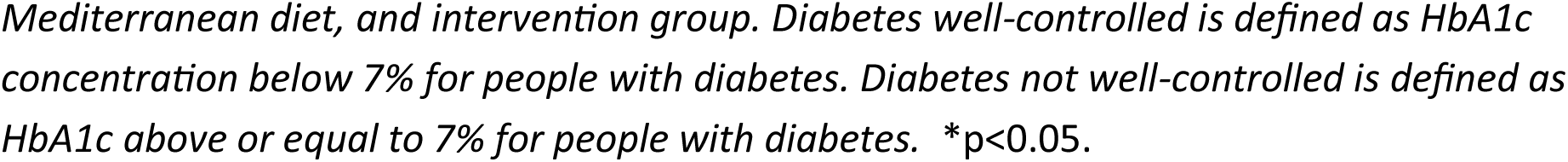
Association of baseline HbA1c concentrations with baseline biomarker concentrations and 5-year change in biomarker concentrations.

| **Biomarker** | **Model** | **Baseline biomarker concentrations**<br>**Overall cohort** | **Baseline biomarker concentrations**<br>**Excluding normoglycemic** | **5-year biomarker change**<br>**Overall cohort $\beta$** | **5-year biomarker change**<br>**Excluding normoglycemic** |
| --- | --- | --- | --- | --- | --- |
| **NT-proBNP** | Model 1 | -0.09 (-0.19, 0.01) | -0.11 (-0.23, 0.01) | 0.00 (-0.07, 0.07) | 0.01 (-0.07, 0.09) |
|  | Model 2 | -0.16 (-0.29, -0.04)* | -0.18 (-0.33, -0.03)* | 0.06 (-0.05, 0.17) | 0.04 (-0.09, 0.17) |
| **hsTnT** | Model 1 | 0.07 (0.03, 0.11)* | 0.09 (0.04, 0.14)* | 0.02 (-0.01, 0.05) | 0.01 (-0.02, 0.05) |
|  | Model 2 | 0.08 (0.02, 0.14)* | 0.10 (0.03, 0.16)* | 0.001 (-0.03, 0.04) | -0.01 (-0.04, 0.03) |
| **hsCRP** | Model 1 | 0.09 (-0.02, 0.19) | 0.08 (-0.04, 0.21) | 0.02 (-0.08, 0.12) | 0.02 (-0.10, 0.14) |
|  | Model 2 | 0.04 (-0.09, 0.17) | 0.03 (-0.12, 0.19) | 0.02 (-0.19, 0.24) | 0.06 (-0.19, 0.30) |
| **PICP** | Model 1 | -0.02 (-0.07, 0.03) | -0.02 (-0.08, 0.04) | 0.01 (-0.04, 0.06) | -0.01 (-0.07, 0.05) |
|  | Model 2 | -0.03 (-0.10, 0.03) | -0.03 (-0.11, 0.04) | 0.03 (-0.07, 0.14) | 0.02 (-0.09, 0.14) |
| **3-NT** | Model 1 | -0.09 (-0.19, 0.01) | -0.11 (-0.22, 0.001) | 0.003 (-0.07, 0.08) | 0.004 (-0.08, 0.09) |
|  | Model 2 | -0.06 (-0.19, 0.06) | -0.09 (-0.23, 0.05) | 0.08 (-0.10, 0.24) | 0.02 (-0.16, 0.20) |

*HbA1c, glycated hemoglobin; NT-proBNP, N-terminal pro-B-type natriuretic peptide; hsTnT, high-sensitivity cardiac troponin T; hsCRP, high-sensitivity C-reactive protein; PICP, procollagen type I C-terminal propeptide; 3-NT, 3-nitrotyrosine; REF, reference group. Model 1: adjusted for age, sex, and education. Model 2: adjusted for age, sex, education, marital status, smoking, physical activity, height, body mass index, depression, and adherence to the energy-reduced*
*Mediterranean diet, and intervention group. Diabetes well-controlled is defined as HbA1c concentration below 7% for people with diabetes. Diabetes not well-controlled is defined as HbA1c above or equal to 7% for people with diabetes. \*p<0.05.*

